# Quantifying the Kinetics of Hematocrit and Platelet Count During Febrile Phase to Develop a Scoring System for Predicting Dengue Shock Syndrome in Adults: A Matched-Case Observational Study from a Hospital in Viet Nam

**DOI:** 10.1101/2025.10.09.25337706

**Authors:** Vu Thi Thanh Mai, Bui Thi Bich Hanh, Ha Vinh, Ho Dang Trung Nghia

**Affiliations:** Pham Ngoc Thach University of Medicine, Ho Chi Minh City, Viet Nam; Hospital for Tropical Diseases, Ho Chi Minh City, Viet Nam

**Keywords:** dengue, warning signs, triage, dengue shock syndrome, dengue shock syndrome score

## Abstract

**Introduction:** Early prediction of dengue shock syndrome (DSS) is crucial for effective patient triage and management. The lack of consensus regarding the precise definition of the laboratory warning sign (WS) -”an increase in hematocrit concurrent with a rapid decrease in platelet count”-has made it difficult to utilize all the WS for predicting DSS.

**Methods:** A matched case observational study was conducted among adult dengue patients hospitalized during the first four days of illness from November 2022 to August 2023, in which each DSS case was matched with three non-DSS ones.

**Results:** There were 448 patients (112 DSS and 336 non-DSS) in this study. An increase in hematocrit concurrent with a rapid decrease in platelet count was observed 1-2 days prior to the development of DSS. The cut-off value of an increase in hematocrit by ≥ 5% concurrent with a decrease in platelet count by ≥ 50% as compared with those of the previous day was found to be predictors of DSS, with a sensitivity of 60.71% and a specificity of 83.04%. A DSS scoring system developed using these two cut-off values, along with the number of clinical warning signs, can be used to predict the risk of DSS in adult patients. It achieved an area under the receiver operating characteristic curve (AUC) of 0.93, sensitivity of 86.6%, and specificity of 87.8%. The Score enables triage of patients into low-, intermediate-, and high-risk groups for appropriate monitoring and management.

**Conclusions:** The warning sign “an increase in hematocrit concurrent with a rapid decrease in platelet count” can be defined as “an increase in hematocrit ≥5% concurrent with a decrease in platelet count ≥50% compared to the previous day”. The DSS score, developed from traditional warning signs, serves as a good predictor of DSS in adult patients.

## INTRODUCTION

Dengue is a mosquito-borne infection caused by dengue virus (DENV). The number of dengue cases reported to the World Health Organization (WHO) has increased from 50,543 cases in 2000 to 14.6 million in 2024 with more than 12,000 dengue-related deaths.^1^ Most cases have been reported in tropical and subtropical areas where the virus is endemic. Dengue cases have also been reported in non-endemic areas, such as mainland Europe, making it a global health concern.^2^ Approximately 1 in 5 infected individuals is symptomatic. The majority of those with symptoms experience a self-limited febrile illness that resolves without complications.. A smaller proportion of patients develop complications leading to life-threatening severe dengue. The WHO Dengue guidelines for diagnosis, treatment, prevention and control, launched in 2009 classified dengue into three categories: dengue without warning signs (WS), dengue with warning signs, and severe dengue.^3^ Severe plasma leakage leading to dengue shock syndrome (DSS) is the most frequent form of severe dengue, accounting for 80% of all cases.^4^ Warning signs are symptoms and signs which occur in the later part of the febrile phase, signaling the possibility of progression to severe dengue. The guidelines list six clinical WSs (abdominal pain or tenderness, persistent vomiting, clinical fluid accumulation, lethargy/restlessness, liver enlargement > 2 cm), and one laboratory WS (an increase in hematocrit concurrent with a rapid decrease in platelet count). While all the clinical WSs are obvious and easy to recognize, the only laboratory WS is just a trend without specific numerical thresholds. How much increase in hematocrit, and how to define rapid decrease of platelet count were not specified. This leads to varied interpretations and practices of this WS among treating physicians.In different studies, an increase in hematocrit has been defined as > 45%, > 46%, >48%, > 50%.^5^ A rapid decrease in platelet count was defined as < 50 × 10^9^/L or < 100 × 10^9^/L in the majority of studies, some used the cut-off values of < 20 × 10^9^/L and even < 150 × 10^9^/L. There is only one study stated clearly the laboratory WS as increase in hematocrit together with a decrease of >10 × 10^9^/L platelets in 24 hours with respect to previous measurement or concurrent with platelet count ≤100 × 10^9^/L.^6^ The lack of unified criteria to define the laboratory WS has made it less useful in daily clinical practice as a predictor of severe dengue.

There are no effective antivirals for dengue; therefore early recognition of complicationsis crucial for ensuring appropriate clinical management. The establishment of reliable systems to triage patients according to their risk of developing severe dengue is a major research priority.^7^ This study was carried out (1) to determine the level of increase in hematocrit and rapid decrease in platelet count between 2 consecutive days that should be considered as a warning sign, and (2) to develop a practical scoring system based in corporation all warning signs to facilitate the stratification of patients into risk categories for progression to dengue shock syndrome, thereby enabling more appropriate clinical management.

## METHODS

### Study design

This is a matched-case observational study.^8^ The study was carried out at a tertiary referral hospital for infectious diseases in Ho Chi Minh city, Viet Nam, from November 2022 to August 2023.

### Inclusion criteria

Patients aged ≥16 years with dengue infection confirmed by a positive NS1 rapid test (Bioline™ Dengue NS1 Ag) or IgM anti-dengue ELISA (MAC-ELISA Dengue, NovaTec), who were admitted to the hospital between day 1 and day 4 of illness and had daily complete blood count monitoring.

### Exclusion criteria

Patients who had undergone anti-shock management for dengue either at the time of admission or at a referring hospital, those with underlying conditions associated with thrombocytopenia or severe anemia, those without at least two complete blood count (CBC) results available from consecutive days prior to the onset of shock, and those who received blood or blood product transfusions during the study period.

### Matching criteria

Each case of DSS was matched to three non-DSS cases based on age (±5 years), sex, and CBCs on the same day of illness, and pregnant status. When more than three potential non-shock controls were available, priority for selection was given to those most comparable in terms of time of hospital admission, day of illness at admission, and admitting ward.

### Recruitment procedure

Two periods included (1) a prospective period from December 2022 to August 2023, and (2) a retrospective period from December 2021 to November 2022, during which data were retrieved from medical records.

All patients were treated in accordance with national 2019 and WHO 2009 guidelines.^9,3^ in which assessments of vital signs and warning signs performed every six hours or more frequently as warranted by the patient’s status, and CBCs obtained on a daily basis. Other biochemical and imaging tests were performed on request of treating physicians as indicated.

### Ethical statement

The study was conducted in accordance with Good Clinical Practice and the guidelines of the Declaration of Helsinki, and was approved by The Ethics Committee for Biomedical Research of The Hospital for Tropical Diseases, Viet Nam (IORG0007145) on 30 November 2022. All patients in the prospective period provided written informed consent. In the retrospective period the data were extracted from medical records so that patients’ consent was waived.

### Definition of variables

- Day of illness: days from fever onset, with day 1 = first day of fever.
- Adult: patients ≥ 16 years old were treated at adult ward.by national policy, so that in this study they were considered as adult.
- Dengue shock syndrome is defined as plasma leakage leading to circulatory failure, manifested by restlessness, irritability or lethargy, cold extremities, rapid and thready pulse, pulse pressure ≤20 mmHg or hypotension including undetectable pulse and blood pressure.^9^
- Hematocrit increase rate (HIR): percentage of Hct increase between 2 consecutive days of illness (D_n_ and D_n-1_) in the febrile phase. HIR (%) = ((Hct_Dn_ - Hct_Dn-1_)/Hct _Dn-1_) X 100.
- Platelet decrease rate (PDR): percentage of platelet decrease between 2 consecutive days in the febrile phase. PDR (%) = ((Plt _Dn-1_ - Plt _Dn_)/Plt _Dn-1_) X 100.
- Body mass index (BMI): using WHO Asia-Pacific standard ≥25 kg/m^2^ for the definition of obesity.

### Outcome measurement

The occurrence of dengue shock syndrome, defined as whether patients progressed to DSS or remained non-shock until discharge.

### Statistical analysis

Data were entered and analyzed using IBM SPSS Statistics version 27 (IBM Corp., New York, USA). In this matched observational study, data from the DSS and non-DSS groups were analyzed using conditional (fixed-effects) logistic regression, with p<0.05 was considered statistically significant.

Regression modeling was performed with conditional stepwise estimation, and diagnostic performance was assessed in terms of sensitivity, specificity, and likelihood ratios. Given the predefined 1:3 matching ratio, the prevalence of DSS in the study cohort was disproportionately high (25%). Consequently, positive likelihood ratios and negative likelihood ratios—measures that are not influenced by prevalence—were used in place of positive and negative predictive values, which are inherently dependent on prevalence. For score development, the final regression model was transformed into a practical scoring system by multiply the coefficients of the adjusted variables by 2 and rounded to get corresponding point values.

## RESULTS

During the study period, a total of 448 dengue patients were enrolled, comprising 112 DSS cases and 336 matched non-DSS controls (Fig. 1).

**Figure 1.**
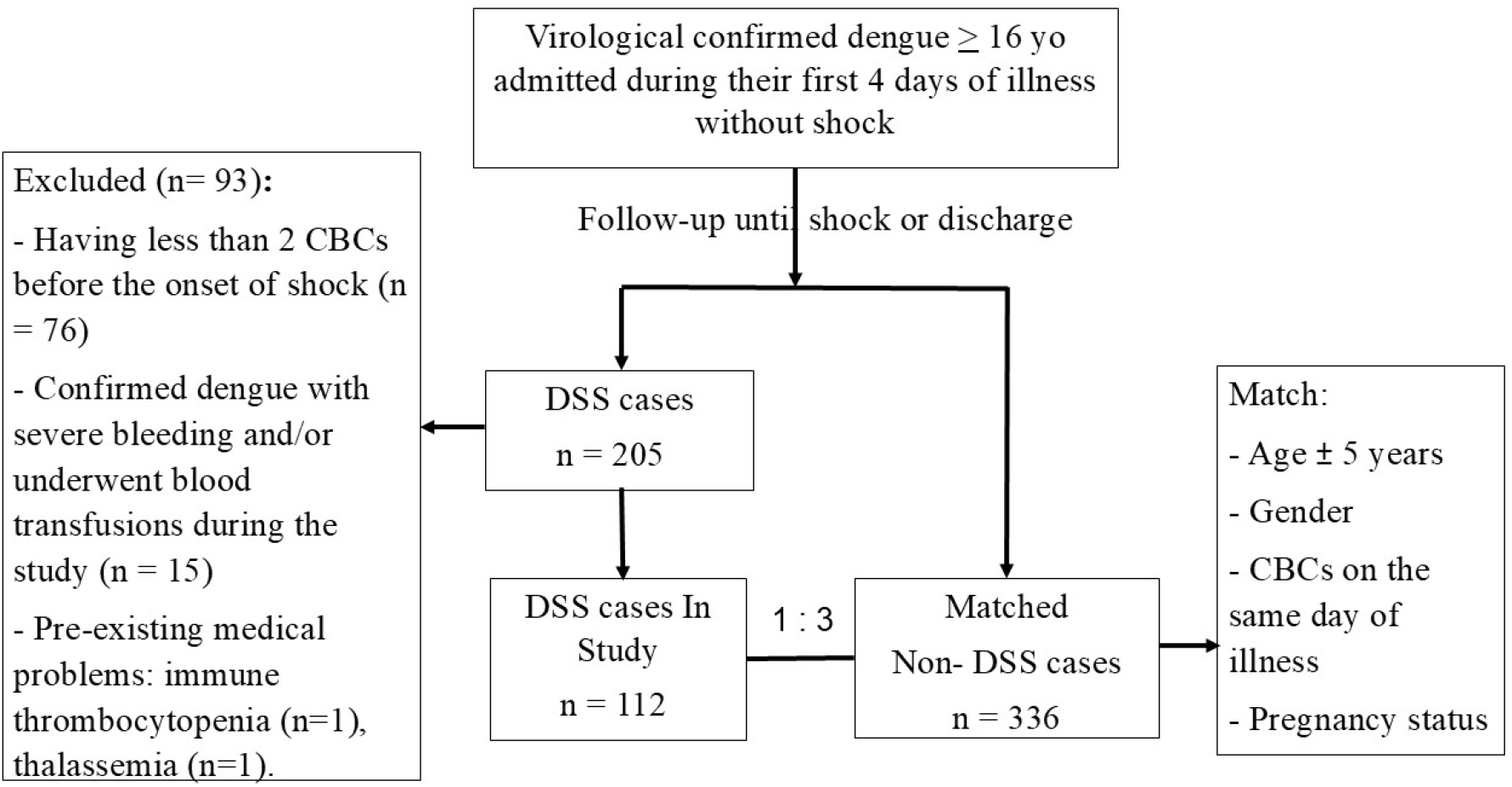
Schema of the recruitment of patients into the study

### Baseline clinical and paraclinical characteristics

The median age of the patients was 25 years, and the male: female ratio was 1.3:1.

Most patients (80%) were admitted on illness day 3 and day 4. Common symptoms included fever (100%), myalgia (72%), persistent vomiting (58%), and abdominal pain/tenderness (50%). Except for fever, these symptoms were significantly more frequent in DSS than non-DSS patients. There was one pregnant patient in the DSS group, who was matched with three pregnant patients in the non-DSS shock dengue group.. Obesity was more prevalent in DSS cases (38% vs. 27%, p = 0.03). Among the 112 DSS patients, shock occurred most often on day 5 (55%), followed by days 6 (28%), day 4 (13%), and day 7 (4%). Clinical or radiological signs of fluid accumulation were also more common in the DSS group (42% vs. 15%, p = 0.001).

Liver transaminase levels were higher in DSS patients than in non-DSS patients, with AST levels exceeding ALT (Table 1).

**Table 1.** Characteristics of the study population.

|  | **DSS**<br>**(N=112)** | **Non-DSS**<br>**(N=336)** | **P-value (\*)** |
| --- | --- | --- | --- |
| Age (years) | 25<br>(21 – 31) | 25<br>(21 – 33) |  |
| Gender male | 64 (57) | 192 (57) |  |
| Pregnancy | 1 | 3 |  |
| Previous dengue | 9 (8) | 17 (5.1) | 0.26 |
| Underlying diseases | 20 (17) | 47 (14) | 0.27 |
| Obesity | 42 (38) | 89 (27) | 0.03 |
| **Clinical characteristics** |  |  |  |
| Fever | 112 (100) | 336 (100) |  |
| Myalgia | 92 (82) | 231 (69) | 0.05 |
| Persistent vomiting | 79 (71) | 179 (53) | 0.001 |
| Abdominal pain | 77 (69) | 144 (43) | 0.001 |
| Hemorrhage (**) | 91 (81) | 152 (45) | 0.001 |
| Petechiae | 71 (63) | 105 (31) | 0.001 |
| Mucosal bleed | 50 (44) | 74 (22) | 0.001 |
| Fluid accumulation | 47 (42) | 49 (15) | 0.001 |
| Number of WS: |  |  |  |
| 0 | 14 (13) | 200 (60) |  |
| 1 | 13 (12) | 87 (26) |  |
| ≥ 2 | 85 (76) | 49 (15) | 0.00001 |
| **Paraclinical characteristics** |  |  |  |
| Transaminase (**)(U/L) |  |  |  |
| AST | 154<br>(109 – 173) | 144<br>(85 – 196) | 0.01 |
| ALT | 107<br>(57 – 183) | 83<br>(57 – 156) | 0.001 |
| Ultrasound of the abdomen (n) | 83 | 249 |  |
| Hepatomegaly | 28 (34) | 39 (16) | 0.001 |
| Thickened gallbladder wall | 49 (59) | 81 (33) | 0.001 |
| Ascites | 43 (52) | 62 (25) | 0.001 |
| Pleural effusion | 28 (34) | 43 (17) | 0.001 |
| Data are n (%) or median (IQR) unless specified. WS: warning sign, DSS: dengue shock syndrome. (*) Conditional logistic regression. (**) AST: Aspartate aminotransferase, ALT: Alanine aminotransferase. |  |  |  |

### Kinetics of Hematocrit and Platelet in DSS Compared with Non-DSS

In non-DSS patients, hematocrit rose slightly from day 2, peaked on day 5, and then declined, whereas in DSS cases it increased more rapidly 1–2 days beforethe onset of shock. Platelet counts declined from day 2 in both groups, with a steeper drop in DSS cases on days 3–5 (Supplementary Figure S1). Compared with non-DSS patients, DSS cases showed significantly higher hematocrit increase rates (HIR) and platelet decrease rates (PDR) from day 3 to day 5, but not from day 2 to day 3 (Figure 2, Figure 3; Supplementary Table S1)

**Figure 2.**
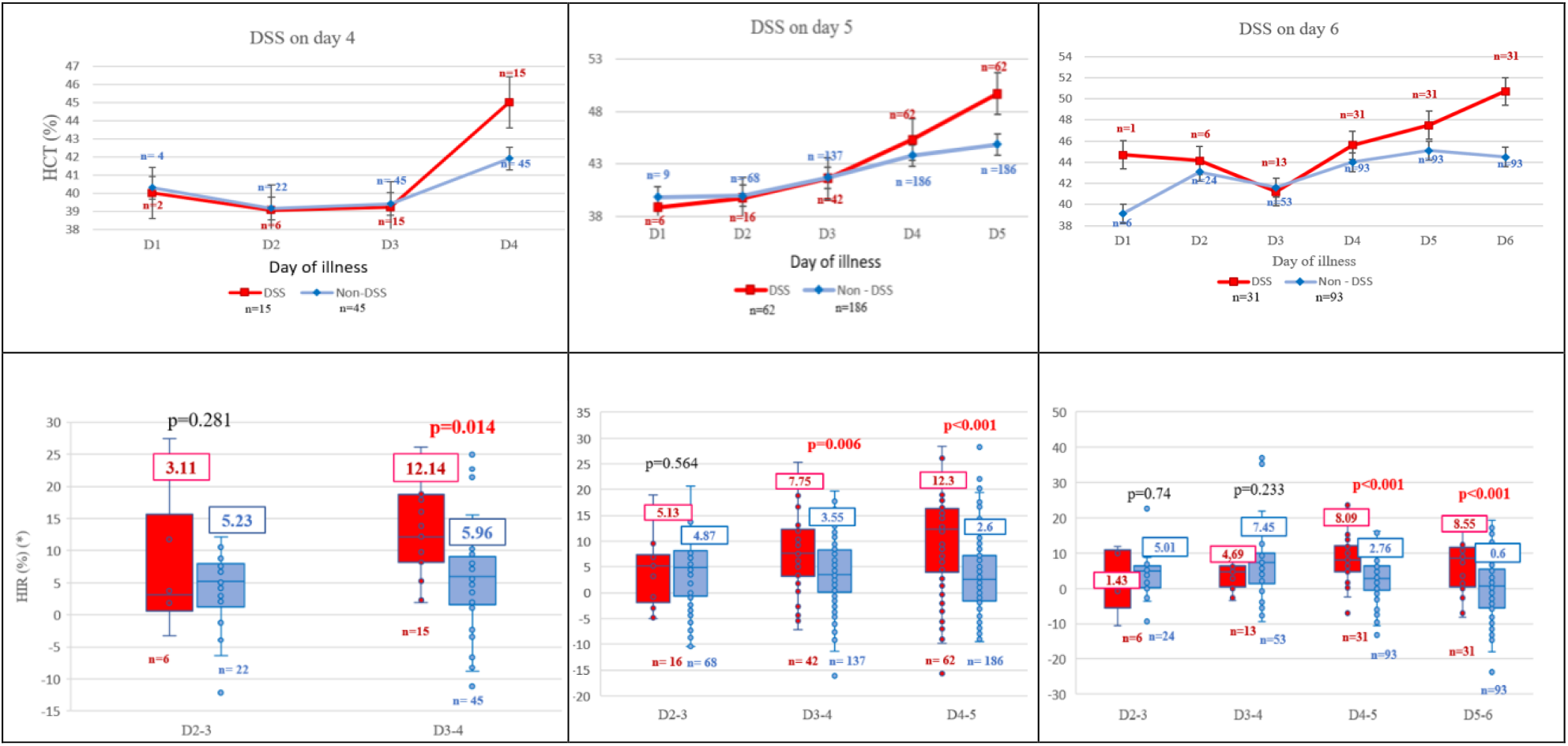
Kinetics of hematocrit (above) and HIR (below) in DSS and non-DSS cases by day of illness at shock onset. (HCT: hematocrit, HIR: hematocrit increase rate, DSS: dengue shock syndrome)

**Figure 3.**
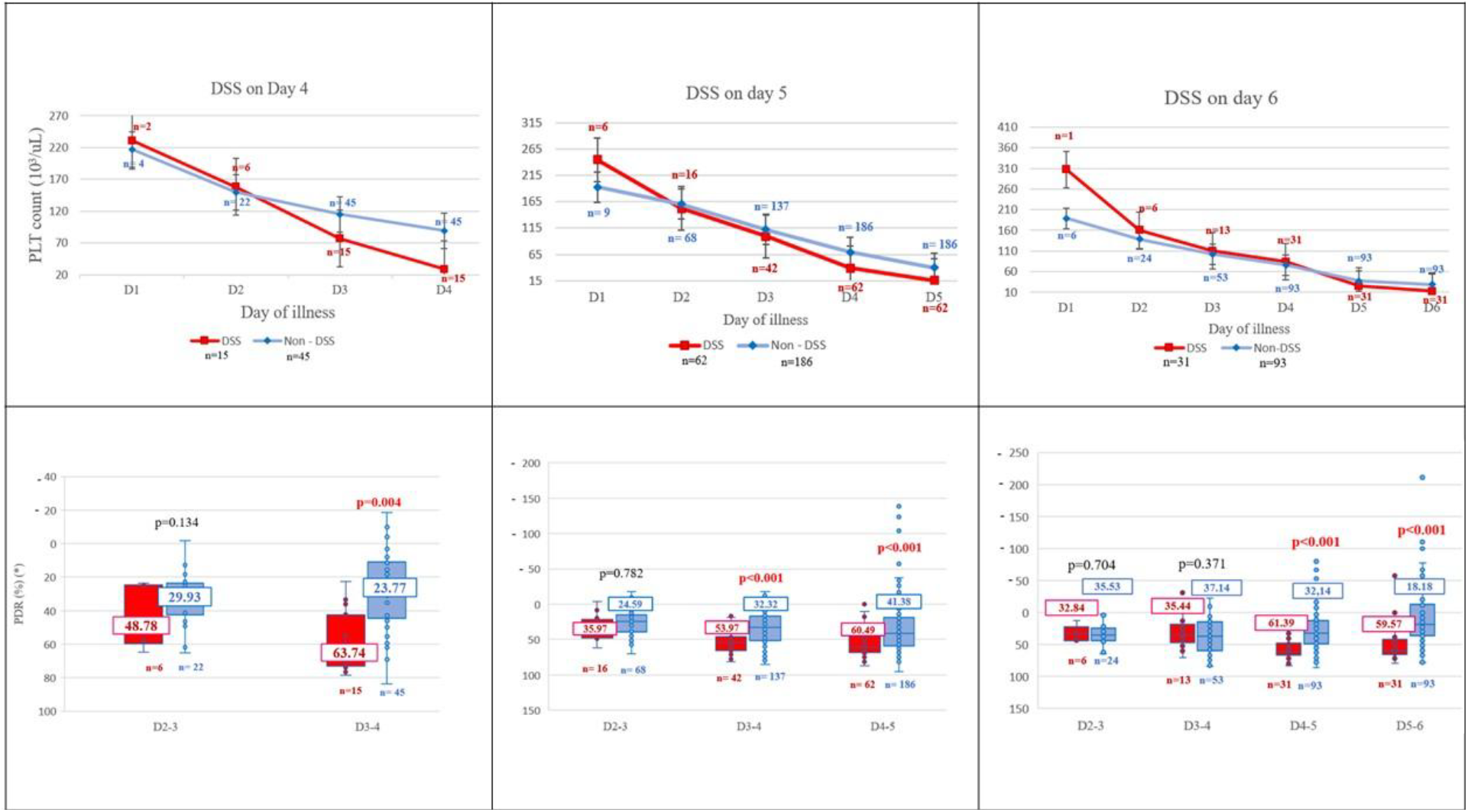
Kinetics of platelet count (above) and PDR (below) in DSS and non-DSS cases by day of illness at shock onset. (PLT: platelet, PDR: platelet decrease rate, DSS: dengue shock syndrome, 10^3^/µL = 10^9^/L)

### Diagnostic value of HIR and PDR in predicting DSS

Based on the Youden index, we have chosen cut-off values of HIR ≥ 5% and PDR ≥ 50%. At these thresholds, the AUCs were 0.79 (95% CI: 0.74–0.84) for HIR and 0.81 (95% CI: 0.78– 0.86) for PDR. Sensitivity, specificity, LR+, and LR– were 78.6%, 63.4%, 2.15, and 0.34 for HIR, and 78.6%, 72.9%, 2.9, and 0.29 for PDR, respectively (See Supplementary Figure S2). The combination of HIR ≥ 5% and PDR ≥ 50% yielded an AUC of 0.81 (95% CI: 0.77–0.85), with sensitivity 69.7%, specificity 83.0%, LR+ 3.6, and LR– 0.47 (Figure 4).

**Figure 4.**
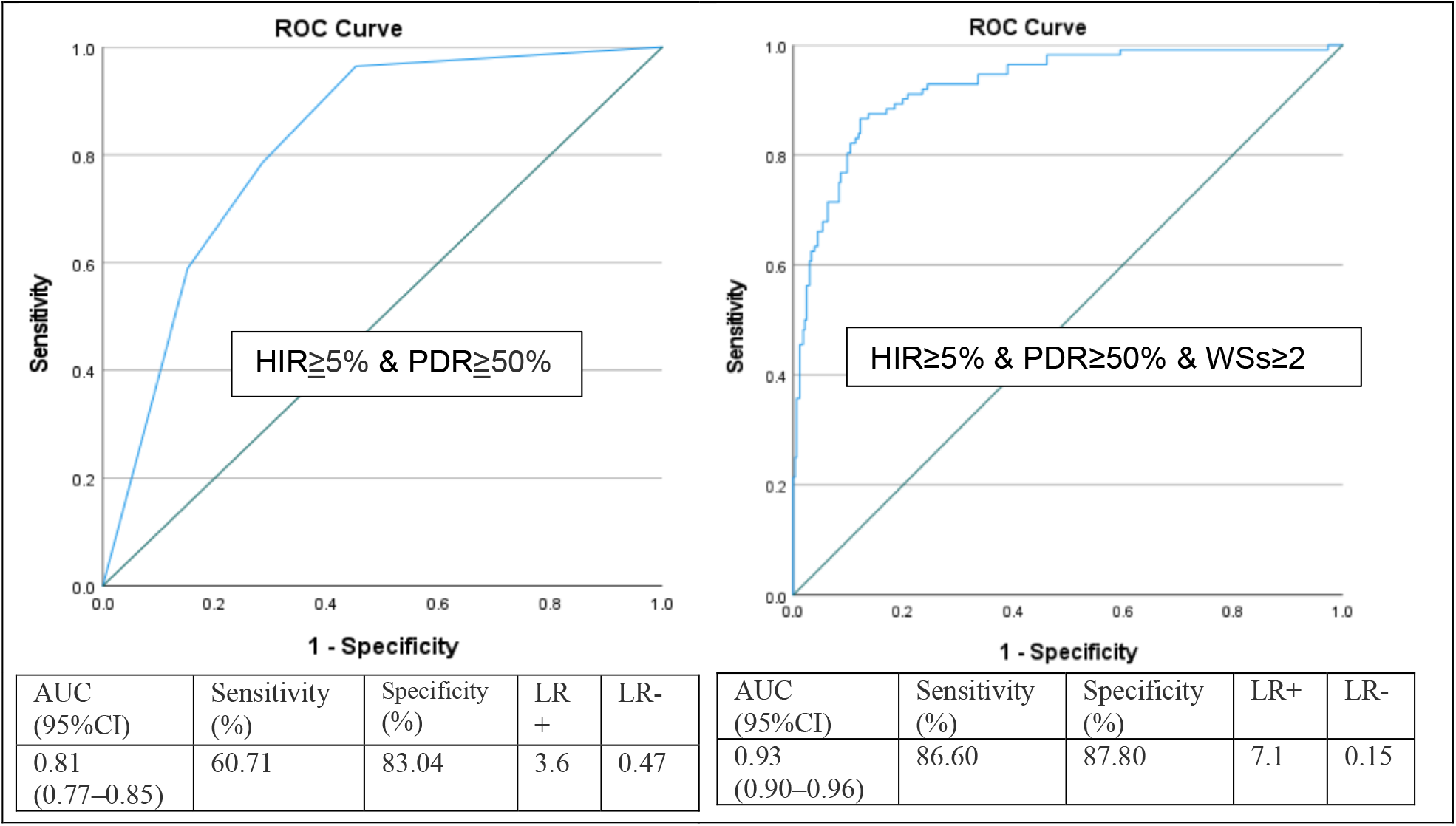
Receiver operating characteristic curve showing the performance of HIR plus PDR (left panel), and of the combination of HIR, PDR and number of clinical WSs (right panel). HIR: hematocrit increase rate, PDR: platelet decrease rate, ROC: receiver operating characteristic curve, AUC: area under the ROC curve, LR: likelihood ratio.

In multivariate analysis, only HIR, PDR, and the number of clinical warning signs remained significantly associated with DSS (Supplementary Table S2). Their combined ROC analysis showed excellent performance, with an AUC of 0.93 (95% CI: 0.90–0.96), sensitivity 86.6%, specificity 87.8%, LR+ 7.1, and LR– 0.15 (Figure 4).

### Prediction Score for Dengue Shock Syndrome

Multivariate regression identified three independent predictors of DSS (p < 0.001): HIR ≥ 5% (coefficient 1.59), PDR ≥ 50% (1.91), and ≥ 2 clinical warning signs (3.06) (Supplementary Table S3). These were converted into a point-based score: 3 points for HIR ≥ 5%, 4 points for PDR ≥ 50%, and 6 points for ≥ 2 WSs. Table 2 showed there were 2 groups at the extremes: a group with LR+ > 10 (DSS score 9, 10, 13) which strongly indicates a high probability of progression to shock, and at the other extreme a group with LR+ at around 2 and LR-< 0.1 (DSS score 3, 4), which means the risk of developing shock is low. So that the risk of DSS based on our scoring system can be stratified into three categories: low risk (≤ 4 points), intermediate risk (6–7 points), and high risk (≥ 9 points). (Table 2).

**Table 2.**
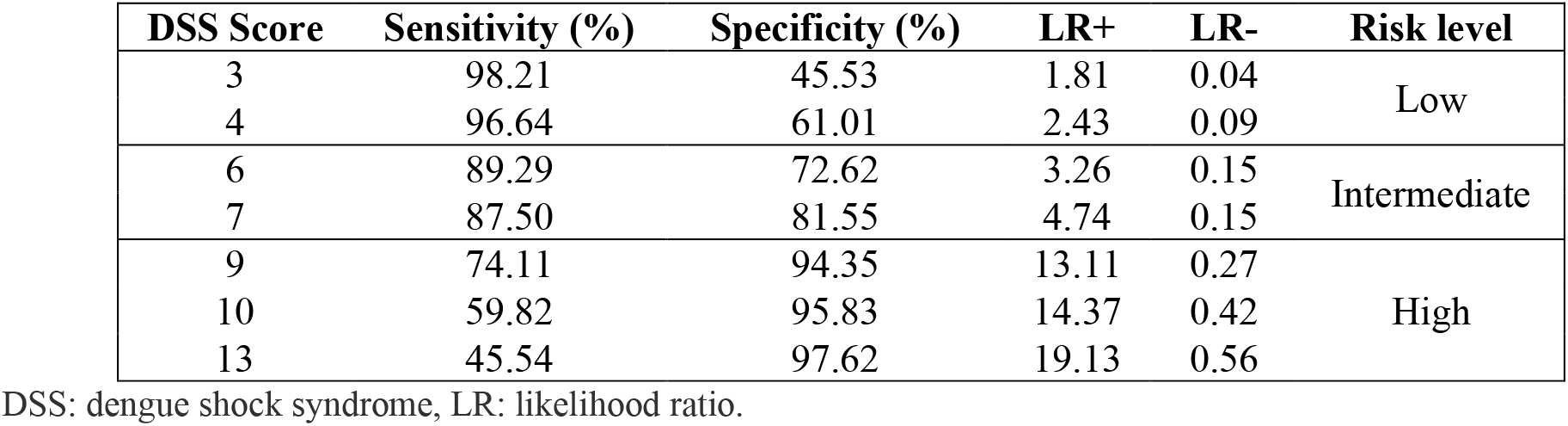
Proposed stratification of DSS risk based on DSS score.

| DSS Score | Sensitivity (%) | Specificity (%) | LR+ | LR- | Risk level |
| --- | --- | --- | --- | --- | --- |
| 3 | 98.21 | 45.53 | 1.81 | 0.04 | Low |
| 4 | 96.64 | 61.01 | 2.43 | 0.09 |  |
| 6 | 89.29 | 72.62 | 3.26 | 0.15 | Intermediate |
| 7 | 87.50 | 81.55 | 4.74 | 0.15 |  |
| 9 | 74.11 | 94.35 | 13.11 | 0.27 | High |
| 10 | 59.82 | 95.83 | 14.37 | 0.42 |  |
| 13 | 45.54 | 97.62 | 19.13 | 0.56 |  |

DSS: dengue shock syndrome, LR: likelihood ratio.

## DISCUSSION

The median age of our patients was 25 years, which was younger than studies reported in studies from Singapore or Taiwan.^10,11^ The median BMI was 24 kg/m^2^ in the DSS group and 22 kg/m^2^ in the non-DSS group (p = 0.07). Fewer than 20% had comorbidities (diabetes, hypertension, or liver disease), and prior dengue infection was rarely reported.

In this matched-case study, most DSS events (95%) occurred between illness days 4 and 6, peaking on day 5. This highlights the need for close monitoring of clinical and laboratory parameters from days 2–3 onward to enable early recognition of signs predicting dengue shock. Hematocrit, reflecting plasma leakage, rose significantly faster in DSS patients compared with non-DSS patients beginning 1–2 days before shock, as captured by the hematocrit increase rate. Thrombocytopenia, a well-recognized feature of dengue, likely results from both reduced bone marrow production and increased peripheral destruction of platelets.^12^ In our cohort, platelet counts began to decline on day 3 and reached their lowest levels on day 6 in both DSS and non-DSS patients, consistent with their proposed role in plasma leakage and inflammation.

The WHO 1997 guidelines defined DHF by a hematocrit increase of ≥20% and a platelet count <100 × 10^9^/L, while WHO 2009 guidelines only considered rising hematocrit concurrent with rapidly falling platelets as a laboratory warning sign,without specifying numerical thresholds. There was one study following the dynamic of hematocrit and platelet count for predicting DSS in children aged 5 to 15 years.^13^ Lam et al. analyzed data from 2,301 patients, of whom 143 developed shock (6%). In a smaller cohort of 908 patients enrolled on day 3 of illness, the researchers used a logistic regression model and graphical analysis to assess the predictive value of daily hematocrit and platelet count for DSS. The study showed that serial platelet counts strongly predicted DSS, whereas daily hematocrit values contributed little.^13^ In our adult population (n=448), HIR and PDR differed significantly between DSS and non-DSS patients 1–2 days before the onset of shock. Our findings are consistent with Lam’s study regarding platelet kinetics but differ in hematocrit performance. This discrepancy may reflect differences in study design: in our study. one case matched with three controls, whereas in Lam’s study, the DSS group was compared with the rest of the study population. The imbalance between case and control may obscure the true positive effect of hematocrit on DSS, which is in smaller magnitude than that of platelet count. Moreover, in graphical analysis, the authors primarily compared absolute values of serial hematocrits in DSS cases versus those in non-DSS cases, while we used dynamic indices (HIR) rather than absolute values. The combination of HIR ≥5% and PDR ≥50% predicted DSS with an AUC of 0.81 (95%CI: 0.77–0.85), sensitivity 60.7%, specificity 83.0%, and positive likelihood ratio 3.6, indicating that patients meeting these cut-offs were 3.6 times more likely to develop shock thanthose who did not. These findings support HIR ≥5% and PDR ≥50% thresholds as practical laboratory warning signs for early identification of dengue patients at risk of DSS.

Many scoring systems have been proposed to predict severe dengue, using either admission clinical and laboratory data or sophisticated biomarkers. Pongban *et al*. developed a scoring system based on six parameters: age, hepatomegaly, hematocrit, systolic blood pressure, white cell count, and platelet count. In their study, 39 out of 90 DSS cases had a score > 11.5 (with scores ranging from 0 to 18).^14^ Based on data from 302 dengue-infected patients in Thailand, Srisuphanunt *et al*. constructed a scoring system for early prediction of severe dengue using six laboratory tests, with a cut-off score of 14 to pedict DSS(score range: 0 to 38.6).^15^ Researchers in Viet Nam used various machine learning models to analyse data from 230 pediatric dengue patients to develop a nomogram for predicting DSS risk^16^. The nomogram using five parameters: albumin, activated partial thromboplastin time, fibrinogen, aspartate aminotransferase, and platelet count, with score ranging from 0 to 350 points. The model performed well in both training and validation sets, with an AUC of 0.985 (95% CI: 0.965-1.000) and accuracy of 0.988 (95% CI: 0.957-0.998) in the training set, and an AUC of 0.945 (95% CI: 0.886-1.000) with an accuracy of 0.951 (95% CI: 0.865-0.989) in the validation set.^16^ The above-mentioned studies required many non-basic laboratory tests, which may not be available at the bedside in endemic areas. A study on adults with dengue in Taiwan used four parameters to develop severity risk score for dengue patients with ≤ 4 days of illness, and two parameters for those with > 4 days of illness. At the cut-off value of 1 point, the sensitivity and specificity were estimated at 70.3% and 90.6% respectively.^17^ The patients in this study had a median age of 51 years in non-severe and 66 years in severe dengue group ,which is much higher than that of our patients. Recently, Madewell and colleagues used various machine learning models to analyze data from dengue cases in Puerto Rico for predicting severe dengue.^18^ Among the 1,708 laboratory-confirmed cases, 415 were classified as severe dengue. Due to an imbalance in the dataset, with non-severe cases being more prevalent, the researchers applied up-sampling to balance the class distribution. The ensemble model using 40 variables achieved the highest overall AUC of 0.977 (sensitivity 95.6%, specificity 93.3%). The performance of each warning sign in predicting severe dengue was also studied using a logistic regression model. The presence of any WSs yielded the highest sensitivity (92.8%) but low specificity (29.2%), with an AUC of 0.611. Combining ≥ 3 WSs resulted in an AUC of 0.713, with sensitivity of 87.2% and specificity of 65.1% (LR+ 2,5, LR-0.37). Despite this high predictive accuracy of machine learning models, the authors acknowledged that their implementation in clinical practice may require computational resources which may not be available in dengue-endemic settings.Therefore, to enhance clinical utility, logistic regression models could compliment machine learning approaches by enabling clinicians to apply these findings more feasibly in practice.^18^ Our study in adults employed daily bedside clinical WSs combined with two laboratory WSs derived from routine CBC results. Multivariate analysis identified three independent predictors of DSS: ≥2 clinical warning signs, HIR ≥5%, and PDR ≥50%. A reduced logistic regression model incorporating these factors showed excellent predictive performance (AUC = 0.93, 95% CI: 0.90–0.96). There are two main differences between the study by Madewel *et al*. and our own that may explain why our approach achieved a higher AUC. First, our study focused only on DSS, while Madewel *et al*. included severe bleeding, a condition in which hematocrit usually decreases rather than increases. This may have interfered with the positive effect of hematocrit increase in predicting DSS. Second, our study included platelete count reduction and fluid accumulation in the WS list, while Madewel *et al*. excluded them.

Based on the sensitivity, specificity, positive and negative likelihood ratios of the DSS score-dependent on the probability of progressing to shockfrom the clinician’s point-of-view-we proposed a practical frameworkto triage dengue patients into three DSS risk groups: low-, intermediate- and high-risk (Table 2).The very low-risk patients (score 0), with a very low probability of developing dengue shock, may be managed as outpatients and be advised to return the following day or sooner if new WSs occur, low-risk patients could be monitored every 6-12 hours, intermediate-risk patients may require monitoring every 3–6 hours; and high-risk patients should have vital signs checked every 30–60 minutes to detect early progression to shock. The score should be re-calculated whenever new WSs or laboratory results become available.

Our study has several limitations. We lacked data on dengue virus serotypes, viral load, and IgG/IgM status, which may influence the risk of shock. However,by using a matching method and recruiting patients during a single epidemic season, the difference between two groups may have been minimized. Although, the sample size was not pre-calculated, our cohort of 112 DSS and 336 non-DSS patients could provide sufficient power for analysis. Requiring at least two consecutive CBCs increased workload and treatment costs, which may limit the applicability of our DSS score in some places. The Pan America Health Organization (PAHO) 2022 ^19^ and WHO 2025 guidelines for clinical management of arboviral diseases ^20^ do not consider thrombocytopenia as a warning sign, since it is not a consequence of extravasation, and therefore not a useful guide for medical professionals in the management of parenteral liquids in dengue.

Our data supported the use of changes in hematocrit and platelet count over time as a warning sign by calculating HIR and PDR to develop a DSS prediction score, not to guide fluid administration. The weakness of our study is that our study is single-centered, and the scoring system has not yet been validated.

Our study has several strengths. First, the dynamic approach to hematocrit and platelet count-by comparing patients’ own hematocrit and platelet count over two consecutive days-depicted individual changes more precisely than comparisions with with population baseline values.

Second, the DSS score is simple and easy to use, eitherthrough mental calculation or via a small spreadsheet compatible with smart devices for rapid bedside risk assessment (See Supplementary file S1, or visit https://sites.google.com/pnt.edu.vn/infectiousdiseases-dss-score to download the DSS score calculator). Thirdly, because dengue with warning signs includes patients with a wide range of severity and outcomes-from mild cases that never progress to shock to critical dengue requiring ICU admission or resulting in death,^21^ our triage schema stratifies patients into three risk levels. This allows re-distributing clinical resources toward high-risk patients while reducing unnecessary workload for low-risk patients. This approach is more practical than the current recommendation of uniformly monitoring vital signs every 2-4 hours for all patients with WS. ^22^

Future research should explore warning signs for other forms of severe dengue including severe hemorrhage, extreme liver enzyme elevations (AST/ALT ≥1000 U/L), and other organ involvement. Identification of specific clinical and laboratory predictors for these outcomes remains an important area for research.

## CONCLUSIONS

We propose that the WHO 2009 laboratory warning sign for dengue could be specified as “an increase in hematocrit ≥5% concurrent with a decrease in platelet count ≥50% compared with those of the previous day.” The DSS Risk Score enables triage of patients into low-, intermediate-, and high-risk groups for appropriate monitoring and management. It may also guide patient selection in future clinical trials of DSS therapies. Further studies in other places, outpatient settings, and across different age groups, including children under 16 years and adults over 60 years old, are needed to validate the score before broader clinical implementation.

## Data Availability

Reasonable data request can be shared through corresponding author

## Authors contributions

Concept and study design: VTTM, BTBH, HDTN, HV

Data acquisition: VTTM

Statistics analysis: VTTM, HDTN

Calculator and formula creation: HV

Writing first draft: HV, VTTM

Draft editing: VTTM, BTBH

Writing final version: HV, HDTN

All authors read and approved the final manuscript, and had final responsibility for the decision to submit it for publication.

## Acknowledgement

We thank all patients for allowing us to use their data for the study. We appreciate all staffs of adult dengue wards for their hardworking caring for the patients. We thank head doctors of adult wards in the hospital for helping in data collecting.

We are particularly in debt to Professor Tran Tinh Hien, Associate Professor Dong Thi Hoai Tam, and Dr. Phan Tu Qui for fruitful comments.

## Disclaimer

The findings and conclusions in this report are those of the authors and do not necessarily reflect the views of Pham Ngoc Thach University of Medicine and The Hospital for Tropical Diseases.

## Conference statement

A preliminary version of the study was presented at The Viet Nam National Scientific Conference on Infectious Diseases and HIV/AIDS, Ha Noi, 31 October - 2 November, 2024. **DOI:** https://doi.org/10.59873/vjid.v3i47

## Conflict of interest

All authors declared no conflict of interest

## Funding

The authors received no financial support for the research and publication of this article.

## Data sharing

Reasonable data request can be shared through corresponding author.

## Notes

### Competing Interest Statement

The authors have declared no competing interest.

### Funding Statement

This study did not receive any funding

### Author Declarations

Ethics committee of the Hospital for Tropical diseases gave ethical approval for this work

### Summary of Updates

- Grammar and spelling corrections - Add a link to website with supplementary materials

