## Supplementary material for "Quantifying the Kinetics of Hematocrit and Platelet Count During Febrile Phase to Develop a Scoring System for Predicting Dengue Shock Syndrome in Adults: A Matched-Case Observational Study from a Hospital in Viet Nam": Kinetics of HIR and PDR in DSS vs Non-DSS: Table S1.docx (from another version of this preprint: version 1, 2025-10-10)

**Table S1.** Kinetics of HIR and PDR in DSS cases according to day first occurrence of shock.

| Kinetics of HIR and PDR of patients had DSS at day 4 versus non-DSS | | | | | | |
| --- | --- | --- | --- | --- | --- | --- |
|  | HIR (%) | |  | PDR (%) | |  |
|  | DSS  (n=15) | Non-DSS  (n=45) | p value | DSS  (n=15) | Non-DSS  (n=45) | p value |
| D2 - D3 | 3.11 | 5.23 | 0.281 | 48.78 | 29.93 | 0.134 |
| D3 - D4 | 12.14 | 5.96 | 0.014 | 63.74 | 23.77 | 0.004 |
| Kinetics of HIR and PDR of patients had DSS at day 5 versus non-DSS | | | | | | |
|  | HIR (%) | |  | PDR (%) | |  |
|  | DSS  (n = 62) | Non-DSS (n =186) | p value | DSS  (n = 62) | Non-DSS (n =186) | p value |
| D2 - D3 | 5.13 | 4.87 | 0.564 | 35.97 | 24.59 | 0.782 |
| D3 - D4 | 7.75 | 3.55 | 0.014 | 53.97 | 32.32 | <0.001 |
| D4 - D5 | 12.3 | 2.6 | < 0.01 | 60.49 | 41.38 | <0.001 |
| Kinetics of HIR and PDR of of patients had DSS at day 6 versus non-DSS | | | | | | |
|  | HIR (%) | |  | PDR (%) | |  |
|  | DSS  (n = 31) | Non-DSS  (n = 93) | p value | DSS  (n = 31) | Non-DSS (n =93) | p value |
| D2 - D3 | 1.43 | 5.01 | 0.740 | 32.84 | 35.53 | 0.704 |
| D3 - D4 | 4.69 | 7.45 | 0.233 | 35.44 | 37.14 | 0.371 |
| D4 - D5 | 8.9 | 2.76 | <0.001 | 61.39 | 32.14 | <0.001 |
| D5 – D6 | 8.55 | 0.06 | <0.001 | 59.57 | 18.18 | <0.001 |
| DSS: dengue shock syndrome, HIR: Hematocrit increase rate, PDR: Platelet decrease rate | | | | | | |
