## Supplementary material for "Quantifying the Kinetics of Hematocrit and Platelet Count During Febrile Phase to Develop a Scoring System for Predicting Dengue Shock Syndrome in Adults: A Matched-Case Observational Study from a Hospital in Viet Nam": Factors associated with DSS: Table S2.docx (from another version of this preprint: version 1, 2025-10-10)

**Table S1.** Factors associated with the development of DSS.

|  | Unadjusted effects | | Adjusted effects | |
| --- | --- | --- | --- | --- |
|  | OR  (95%CI) | *P* ^(*)^ | OR  (95%CI) | *P* ^(*)^ |
| Previous dengue | 1.59  (0.70 – 3.56) | 0.262 |  |  |
| Obesity | 1.67  (1.07 – 2.59) | 0.024 | 1.14  (0.77 – 1.69) | 0.505 |
| Number of clinical warning signs | 3.64  (2.65 – 4.99) | <0.001 | 3.04  (2.03 – 4.56) | <0.001 |
| HIR | 1.18  (1.13 – 1.23) | <0.001 | 1.14  (1.07 – 1.22) | <0.001 |
| PDR | 1.06  (1.04 – 1.07) | <0.001 | 1.04  (1.02 – 1.06) | <0.001 |

DSS: dengue shock syndrome, HIR: hematocrit increase rate, PDR: platelet decrease rate
