## Supplementary material for "Quantifying the Kinetics of Hematocrit and Platelet Count During Febrile Phase to Develop a Scoring System for Predicting Dengue Shock Syndrome in Adults: A Matched-Case Observational Study from a Hospital in Viet Nam": DSS score: Table S3.docx (from another version of this preprint: version 1, 2025-10-10)

|  | Coefficient (*) | *P* | OR | 95%CI | DSS Score point |
| --- | --- | --- | --- | --- | --- |
| HIR ≥ 5% | 1.59 | < 0.001 | 4.91 | 2.21 – 10.88 | 3 |
| PDR ≥ 50% | 1.91 | < 0.001 | 6.74 | 3.06 – 14.88 | 4 |
| Number of clinical WS ≥ 2 | 3.06 | < 0.001 | 21.28 | 8.82 – 51.35 | 6 |

(*) Using conditional stepwise regression

OR: odd ratio, DSS: dengue shock syndrome, HIR: Hematocrit increase rate, PDR: Platelet decrease rate, WS: warning sign.
