## Supplementary material for "Quantifying the Kinetics of Hematocrit and Platelet Count During Febrile Phase to Develop a Scoring System for Predicting Dengue Shock Syndrome in Adults: A Matched-Case Observational Study from a Hospital in Viet Nam": Link to DSS score calculator: DSS Score Calculator_Link to download.docx (from another version of this preprint: version 1, 2025-10-10)

Link to download DSS Score Calculator

<https://docs.google.com/spreadsheets/d/19KmTyHTG0ly3CXAd97t2eQsI-dn4F2LW/edit?usp=sharing&ouid=100670763844769408233&rtpof=true&sd=true>

https://pnteduvn-my.sharepoint.com/:x:/g/personal/vinhha_pnt_edu_vn/EcBLCN3dqSdHgKX1WEierCcBDM0bz5umhMeLA5a3udk_5A?e=orM8cp&nav=MTVfezk0RDQwMjlGLTM5RkYtNDI2Qy1BOTBFLTdFODI1NEZGMkE3Nn0
